# Wastewater-based epidemiology for tracking COVID-19 trend and variants of concern in Ohio, United States

**DOI:** 10.1101/2021.06.08.21258421

**Authors:** Yuehan Ai, Angela Davis, Danial Jones, Stanley Lemeshow, Huolin Tu, Fan He, Peng Ru, Xiaokang Pan, Zuzana Bohrerova, Jiyoung Lee

## Abstract

The global pandemic caused by severe acute respiratory syndrome coronavirus 2 (SARS-CoV-2) has resulted in more than 129 million confirm cases. Many health authorities around the world have implemented wastewater-based epidemiology as a rapid and complementary tool for the COVID-19 surveillance system and more recently for variants of concern emergence tracking. In this study, three SARS-CoV-2 target genes (N1, N2, and E) were quantified from wastewater influent samples (n = 250) obtained from the capital city and 7 other cities in various size in central Ohio from July 2020 to January 2021. To determine human-specific fecal strength in wastewater samples more accurately, two human fecal viruses (PMMoV and crAssphage) were quantified to normalize the SARS-CoV-2 gene concentrations in wastewater. To estimate the trend of new case numbers from SARS-CoV-2 gene levels, different statistical models were built and evaluated. From the longitudinal data, SARS-CoV-2 gene concentrations in wastewater strongly correlated with daily new confirmed COVID-19 cases (average Spearman’s *r* = 0.70, *p* < 0.05), with the N2 gene being the best predictor of the trend of confirmed cases. Moreover, average daily case numbers can help reduce the noise and variation from the clinical data. Among the models tested, the quadratic polynomial model performed best in correlating and predicting COVID-19 cases from the wastewater surveillance data, which can be used to track the effectiveness of vaccination in the later stage of the pandemic. Interestingly, neither of the normalization methods using PMMoV or crAssphage significantly enhanced the correlation with new case numbers, nor improved the estimation models. Whole-genome sequencing result showed that those detected SARS-CoV-2 variants of concern from the wastewater matched with the clinical isolates from the communities. The findings from this study suggest that wastewater surveillance is effective in COVID-19 trend tracking and variant emergence and transmission within a community.

## 1. Introduction

Coronavirus disease 2019 (COVID-19), first reported in Wuhan, China, is caused by Severe Acute Respiratory Syndrome Coronavirus 2 (SARS-CoV-2) and has resulted in more than 2.8 million deaths and 129 million confirmed cases globally as of April 2021 (1-2). Community and intrafamily transmission is one of the most common modes of human-to-human transmission of SARS-CoV-2 (3). However, the transmission of COVID-19 can occur before the confirmation of a clinical diagnosis (4). For more effective control and timely monitoring of the outbreaks, wastewater-based SARS-CoV-2 surveillance has been implemented as a complementary tool for the COVID-19 surveillance system (5, 6). Wastewater-based epidemiology (WBE) has been employed to monitor a variety of pathogenic viruses around the world such as Polio, Dengue, Norovirus, and SARs-CoV (7). WBE targets the DNA/RNA residue from viruses, which serve as a population biomarker of the pathogen. Not like other respiratory viruses, SARS-CoV-2 is found in the gastrointestinal tracts and stools of majority of infected people (8, 9). The discharged virus has been detected in wastewater streams at the early stages of the pandemic and even before the first recorded case (10-13). With the analysis of viral signals in population-pooled wastewater samples, WBE can provide early warnings of COVID-19 emergence at a community level (14-17).

The WBE surveillance of COVID-19 is advantageous in many other ways. Firstly, with approximately 105,600 wastewater treatment plants operating globally, 27% of the global population can benefit from health information provided by WBE (5). Wastewater can capture signals from symptomatic as well as pre-symptomatic/asymptomatic carriers, which tend to be under-detected by clinical tests. Secondly, wastewater provides a longer detection window for the SARS-CoV-2 carriers since RNA signals in fecal samples showed longer persistency than in oropharyngeal swabs (9). Thirdly, it is hard to gain a “real-time” picture of the pandemic from clinical screening due to backlogs of test results up to 10 days (18-19). WBE is capable of generating results in a real-time manner with a relatively low cost compared to individual clinical testing, enabling the decision-makers to identify outbreak hotspots and take timely actions (20). Moreover, WBE can aid in monitoring the epidemic progression by giving reliable information on the effectiveness of intervention strategies. As several SARS-CoV-2 variants have emerged, some studies successfully employed WBE to investigate the circulating viral variants in the wastewater through high-throughput sequencing (21-23). Therefore, WBE has been adopted as a COVID-19 trend tracker and more recently for detecting variants by public health authorities in the United States.

At present, most wastewater-associated studies have focused on municipal wastewater and covered a relatively short period of time at the early stages of the pandemic. In an effort to help with the long-term monitoring of the spread of COVID-19 across the state of Ohio, the Ohio Department of Health (ODH), the Ohio Environmental Protection Agency (Ohio EPA), and Ohio Water Resources Center (Ohio WRC) at The Ohio State University established the Ohio Coronavirus Wastewater Monitoring Network (6) using participating laboratories in Ohio. The present study contributes to the network by generating critical wastewater-based information for the populations in nine various sewage catchments in central Ohio, including the largest city, Columbus, and other urban and rural areas. A quantitative method was developed and validated for the measurement of SARS-CoV-2 gene targets in wastewater samples. To further evaluate the feasibility of using WBE as a predicting/modeling tool for the COVID-19 outbreak dynamics, correlations between the concentrations of SARS-CoV-2 genes in wastewater and the clinical COVID-19 case number in their corresponding sewershed areas were investigated. In order to compensate for the fluctuation in fecal material caused by dilution, we investigated normalization methods with two of the most prevalent human fecal viral indicators, pepper mild mottle virus (PMMoV) and cross-assembly phage (crAssphage) (22, 24-28). Moreover, this study explored whether the wastewater matrix can serve as a sentinel piece for detecting SARS-CoV-2 variants of concern within a community.

## 2. Materials and Methods

### 2.1. Sampling sites and wastewater collection

Nine wastewater treatment plants (WWTPs) in central Ohio were involved in this study. Two of the WWTPs (Jackson Pike and Southerly) serve different catchments in Columbus, which is the largest city in Ohio with a population of around 900,000 (29). The sewersheds of the other wastewater facilities cover 7 smaller Ohio cities (Athens, Circleville, Lancaster, Marietta, Marysville, Newark, and Zanesville) in urban and rural areas with population ranges from 14,000 to 49,000 (29). Confirmed COVID-19 case numbers and the boundaries of all 9 sewershed catchments vary (Figure 1), and details on the serving population and operating characteristics of the WWTPs are summarized (Table 1). Approximately one liter of 24-hour composite samples were collected from the WWTPs twice a week, representing the untreated wastewater influent of Sunday and Tuesday. The sampling period started in late July 2020, with varied starting dates among sites, and ended at the first week of January 2021. Wastewater samples obtained from Jackson Pike, Southerly, and Newark WWTPs were delivered to the lab and processed on the sampling day. Samples from the other utilities were shipped on ice overnight and processed on the following day. Samples were immediately stored at 4°C until further processing. Samples that were delayed in their shipment and subjected to temperature abuse (> 10°C) were not processed.

**Figure 1.**
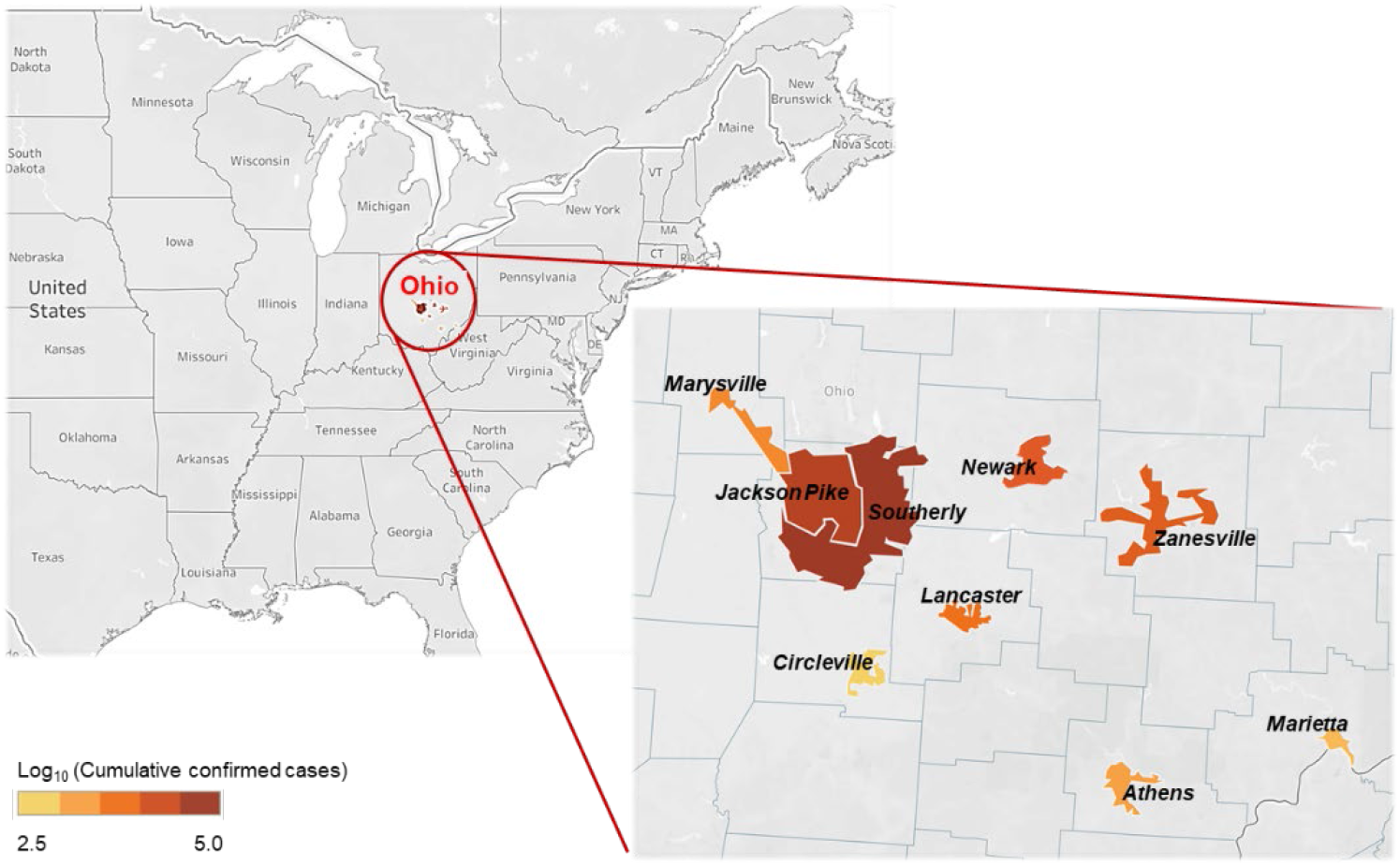
Geographic boundaries and locations of nine sewersheds and cumulative confirmed COVID-19 case numbers.

**Table 1.** Summary of WWTP operating characteristics and each sewershed population with confirmed case numbers during the study period.

| WWTP name | City | County | Sewer type | Average flowrate (MGD) | Population served | Cumulative confirmed cases<sup>a</sup> | Cumulative incidence (cases /100,000 residents) |
| --- | --- | --- | --- | --- | --- | --- | --- |
| Jackson Pike | Columbus | Franklin | Combined | 82 | 645,940 | 41,557 | 6,434 |
| Southerly | Columbus | Franklin | Combined | 130.4 | 654,817 | 46,248 | 7,063 |
| Athens | Athens | Athens | Separate | 2.6 | 24,536 | 1,523 | 6,207 |
| Circleville | Circleville | Pickaway | Separate | 2.01 | 13,965 | 692 | 4,955 |
| Lancaster | Lancaster | Fairfield | Combined | 7.67 | 24,303 | 2,616 | 10,764 |
| Marysville | Marysville | Union | Separate | 4.37 | 24,677 | 2,166 | 8,777 |
| Marietta | Marietta | Washington | Separate | 2.84 | 15,284 | 842 | 5,509 |
| Newark | Newark | Licking | Combined | 9 | 45,000 | 2,889 | 6,420 |

| Zanesville | Zanesville | Muskingum | Combined | 5.8 | 47,500 | 2,653 | 5,585 |
| --- | --- | --- | --- | --- | --- | --- | --- |
| <sup>a</sup> Cumulative confirmed cases from the beginning of the pandemic to 01/02/2021. |  |  |  |  |  |  |  |

### 2.2. Wastewater processing: Virus filtration and concentration

To concentrate virus from wastewater samples, two approaches were used after optimization. Initially, an adsorption-precipitation-based method was adopted. Virus was first adsorbed and eluted from a positively charged filter unit (ViroCap, Scientific Methods, Inc., Granger, IN, USA), followed by concentrating via organic flocculation and centrifugal ultrafiltration (Supplementary Method S1) (30). For faster turnaround, from the second month of the study, we employed a protocol consisting of solid removal and viral concentration. Each wastewater sample was processed in duplicate. First, 100 mL of raw wastewater with 0.05% Tween-20 were centrifuged at 4°C, 2,500 x g for 10 minutes for large solid removal. Small debris and bacteria were further removed by filtering the supernatant using a 0.45 μM sterile filter unit (Milipore, Burlington, MA, USA). Then, the filtrate was concentrated using a 0.05 μm Hollow Fiber Polysulfone Concentrating Pipette Select tips (Innovaprep, Drexel, MO, USA). Approximately 200 μL of viral concentrate was then eluted following the manufacturer’s instruction with some modifications: valve open for 600 ms, 1 pulse, foam factor of 10, valve start time of 3.0 seconds, flow end of 10 seconds, flow minimum start time of 40 seconds, delay of 3.0 seconds, pump at 25%, pump delay time of 1 second, and stored at -80°C for downstream analysis. Recovery efficiency of the method was evaluated by spiking (∼10^9^ gene copies/mL) with three different surrogates: male specific coliphage MS2 (ATCC cat. No. 15597-B1), bovine coronavirus (BCoV strain ML-6 mebus), and human coronavirus OC43 (ATCC cat. No. VR-1558) (32-33).

### 2.3. RNA/DNA extraction and RT-ddPCR analysis

RNA/DNA was extracted from the viral concentrate using an RNeasy PowerMicrobiome Kit (QIAGEN, Germantown, MD, USA). Reverse transcription was conducted with the High-Capacity cDNA Reverse Transcription Kit (Applied Biosystems, Pittsburgh, PA, USA). 3 μL cDNA was applied to the quantification of SARS-CoV-2 genome equivalents using a droplet digital PCR (ddPCR) platform (Bio-rad QX200 system). Three ddPCR assays were developed to target two nucleocapsid (N) genes and the envelope (E) gene of SARS-CoV-2. The N gene assay employs two primers/probe sets from U.S. Centers for Disease Control and Prevention (CDC), amplifying the N1 and N2 regions (34-35). The E gene assay is based on the E_Sarbeco primers/probe set recommended by WHO (36). Quantifications for three SARS-CoV-2 surrogates (MS2, BCoV, and OC32) and two human fecal indicators (PMMoV and crAssphage) were also conducted with diluted cDNA (37-41). The reaction mixture (20 μL) contains ddPCR supermix for probes (Bio-Rad, Hercules, CA, USA), DNase-& RNase-free water, 900 nM of forward and reverse primers, 250 nM of probe, and templates. Firefly (*Coleoptera*) Luciferase control RNA (Promega, Madison, WI, USA) was implemented as an internal amplification control for the detection of PCR inhibition (42). ddPCR mixture with or without wastewater cDNA template were spiked with an equal titer of Luciferase cDNA. PCR inhibition was assessed by comparing the difference in Luciferase gene amplification. Primer/probe sequences and ddPCR parameters used in this study are summarized (Table S1). Briefly, after droplet generation using the QX200 Droplet Generator, target genes were amplified with a Bio-Rad C1000 Touch Thermal Cycler. The cycling conditions included an initial denaturing step at 94°C for 10 minutes, followed by 40-45 cycles of 94°C for 30 seconds and annealing for 60 seconds. A final incubation step was performed at 98°C for 10 minutes and then a final hold of 4°C. After amplification, gene concentrations were quantified using a QX200 droplet reader and QuantaSoft (V 1.7; Bio-Rad). Two technical replicates were performed for each ddPCR assay.

### 2.4. Quantification and statistical analysis

COVID-19 case numbers were retrieved from ODH COVID-19 Dashboard (6), reflecting the confirmed case counts by the estimated symptom onset date. The boundaries of the sewersheds were mapped with Tableau (V 2020.1) (Figure 1). 3-day, 5-day, and 7-day moving averages were calculated from the case numbers using “zoo” package in RStudio (V 1.3.1093). All other plots were generated with “ggplot” package in Rstudio. Target gene copy numbers were calculated as the mean of four replicates (two biological and two technical replicates). The limit of quantification (LOQ) of ddPCR assay is two gene copies/reaction. For detectable-but-not-quantifiable (DNQ) measurements, the results are recorded as one-half of the LOQ. All statistical analyses were performed in RStudio using “ggpubr”, “stats”, and “rstatix” packages. A *p-value* < 0.05 is considered statistically significant. The strength of a linear association was first assessed through a Pearson correlation coefficient. All essential assumptions were examined (normality and linearity). SARS-CoV-2 RNA concentrations and case counts were fitted into a linear model. Spearman’s non-parametric correlation coefficients were also computed since they were less dependent on the underlying distributions being normal. Additionally, polynomial regressions models were fit. Significance of regression models was assessed via the F-test.

### 2.5. SARS-CoV-2 genome sequencing and mutation analysis

A subset of samples of early January 2021 was selected for next-generation sequencing (NGS) by hybridization and/or amplicon methods. For probe-bait NGS, extracted RNA underwent 1^st^ and 2^nd^ strand cDNA synthesis (NEBNext® Ultra# II Non-Directional RNA Second Strand Synthesis Module, NEB, Ipswich, MA). Library construction was performed with KAPA Hyper Prep with KAPA HiFi HotStart Library Amplification Kit (Roche Diagnostics, Indianapolis, IN), with subsequent hybridization with SARS-CoV-2 bait probes (IDT, Coralville, CA) and were then sequenced on NextSeq (Illumina, San Diego, CA). Amplicon sequencing was performed using the CovidSeq kit and sequenced on NextSeq. Analysis tools include custom pipelines utilizing GATK and Mutect2 (Broad Institute) and Dragen SARS-COVID variant detection (Illumina). Viral sequences were strain-typed using Pangolin (43) and NextStrain criteria (44).

For comparison with wastewater findings, sequencing of nasal or nasopharyngeal swab extracted RNA from SARS-CoV-2 positive PCR samples from The Ohio State University Wexner Medical Center was performed. The laboratory received samples from sites across central Ohio, including all sites in this survey except Athens and Marietta. NGS was performed with CovidSeq as above or using the AmpliSeq SARS-CoV-2 Research Panel on Ion Chef-S5 instruments (Thermofisher, Waltham, MA). The lower limit of detection of a mutation for the amplicon methods was approximately 5% variant fraction at a mean depth of 200-1000 reads.

## 3. Results and discussion

### 3.1. Performance of the wastewater concentrating and detection methods

A variety of methods have been adopted around the world to concentrate and quantify SARS-CoV-2 from wastewater. The reliability, reproducibility, and sensitivity of these methods needs to be validated to make better use of wastewater data (45). The performance of the two viral filtration and concentration methods in this study was evaluated by monthly recovery efficiency tests with three SARS-CoV-2 surrogates. MS2 is a non-enveloped bacteriophage widely used as a surrogate for viral pathogens (46-47). For a better understanding of the efficiency of the methods on SARs-CoV-2, two other enveloped coronaviruses (BCoV and OC43) were used in this study. The Concentrating Pipette (CP)-based concentration method was more effective than the ViroCap-based method, especially in recovery efficiency and speed. The time needed for the CP protocol was ten times shorter than the ViroCap-based method. The recovery efficiency of MS2 with the CP method (53.6%) was two times higher than that with the ViroCap-based method (24.7%). ViroCap was less effective in recovering enveloped coronaviruses from wastewater (BCoV: 7.2%). The recovery efficiency of BCoV with the CP-based method varied among the samples tested (ranged from 16.8% to 53.2%), indicating that the efficiency may be dependent on the characteristics of the wastewater matrix, such as the solid contends (48). To enhance the recovery efficiency and shorten the processing time, we switched to the CP-based protocol after the first month. One potential concern of this rapid CP method was that solids are removed prior to virus concentration. Since a previous study found that enveloped viruses tend to be more associated with the surface of solids in wastewater than non-enveloped viruses (49), the partitioning of the coronavirus surrogates in sample fractions was investigated for our methods. In the spiking test, only <0.2% of the BCoV was recovered from the pellet after centrifugation. Compared to the spiked BCoV, SARS-CoV-2 probably had longer residence time in wastewater, thus, higher percentage of SARS-CoV-2 (∼10%) was found in the pellet portion. Since the majority (∼90%) of SARS-CoV-2 RNA signal was detected in the viral concentrate, the virus in the solid (pellet) portion of wastewater was not included in this study.

For accurate quantification of SARS-CoV-2 gene concentration, presence of PCR inhibition in the samples should be checked. In our study, no PCR Inhibition was detected. It might be due to two reasons: the Qiagen kit used for RNA extraction includes several inhibitor removal steps; and ddPCR is more robust in handling inhibition-prone environmental samples than conventional quantitative PCR (50).

### 3.2. SARS-CoV-2 gene concentrations in wastewater

During this 5-month study, three SARS-CoV-2 genes (N1, N2, and E gene) were detected in all 250 wastewater influents, with concentrations ranging from 1 × 10^2^ to 1 × 10^5^ gene copies/L of wastewater. The overall observed trend was that the SARS-CoV-2 gene concentrations stayed relatively stable at the initial stages of the study, increased rapidly in mid-October, peaked in late-November, and plateaued in December. This trend agrees with the COVID-19 daily new confirmed case trend seen in Ohio (51).

In general, wastewater from Jackson Pike and Southerly, the two Columbus WWTPs, showed 1-15 times higher viral concentrations than that from the other utilities in smaller cities (Figure 2). By the end of 2020, Columbus had about 15-60 times more confirmed COVID-19 cases than the other 7 cities, while the cumulative case incidence was at the same magnitude among all the sites (5,000-10,000 cases per million residents, Table 1). It is important to note that the variation of SARS-CoV-2 RNA concentration in wastewater among the sewersheds was not proportional to the variation in confirmed cases nor incidence. This finding suggests that wastewater is a complex matrix due to the variation in many factors, such as individual viral shedding amount and duration, RNA degradation rates, and migration of carriers. A previous study proposed an estimation of the prevalence of COVID-19 infection within a catchment from SARS-CoV-2 gene copies in wastewater (14). Their model embedded other parameters of high uncertainty and variability, including flow rate and per capita production of wastewater, as well as viral shedding rate (14). Despite these potential uncertainties, we support that WBE is still a powerful tool in capturing the real-time infection trend in communities.

**Figure 2.**
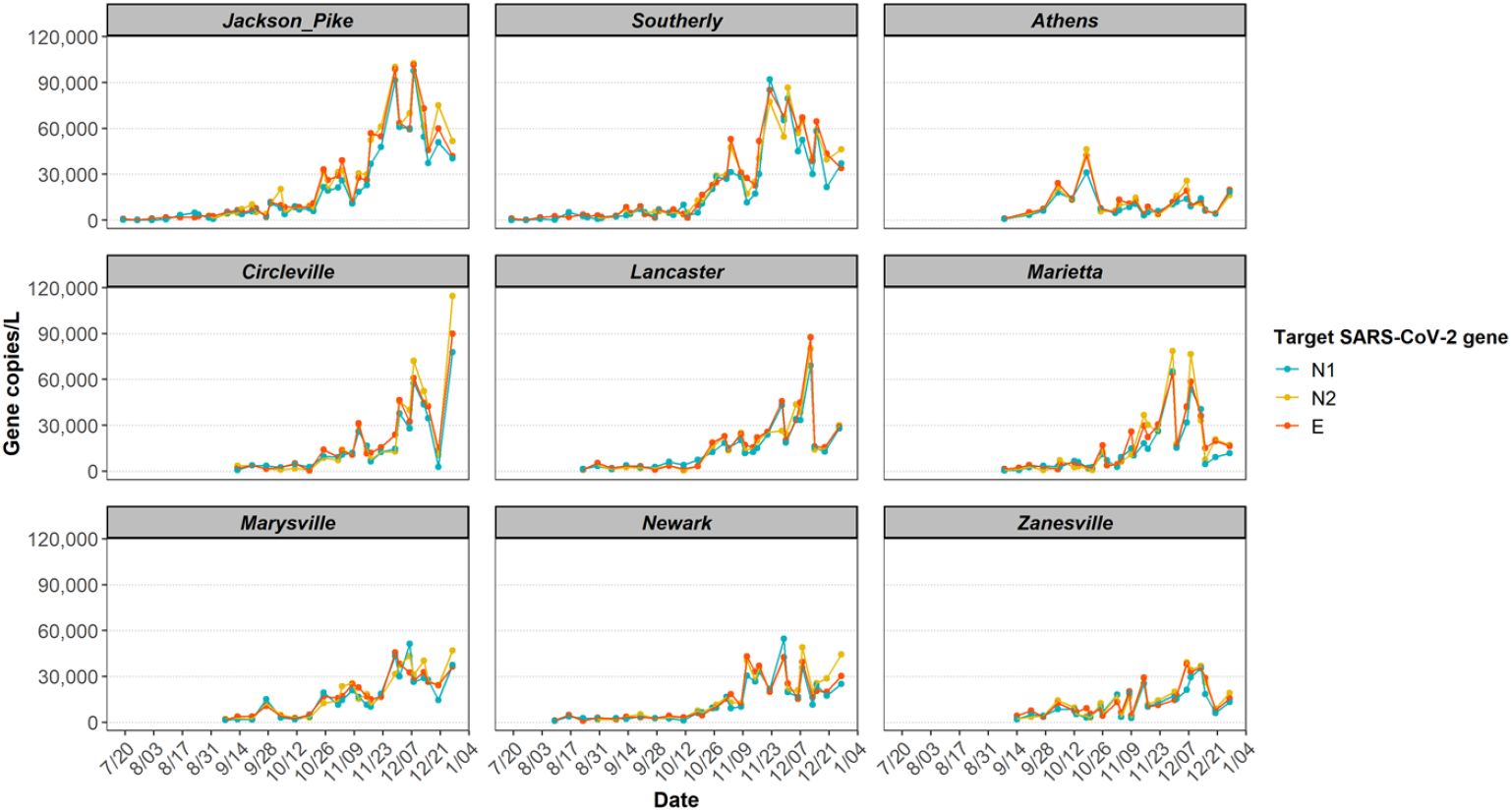
Longitudinal SARS-CoV-2 concentration trend in wastewater measured by N1, N2 and E genes from 9 wastewater catchments in Ohio in 2020.

### 3.3. Correlations between wastewater SARS-CoV-2 concentrations and COVID-19 cases

It is notable that the increased concentration of SARS-CoV-2 genes in wastewater during November and December 2020 coincided with the post-holiday COVID-19 surge, resulting from increased family gathering and travel. This result shows the usefulness of WBE in pinpointing epidemic hotspots (5). We hypothesized that the level of SARS-CoV-2 genes in wastewater correlate with the COVID-19 confirmed cases, so regression analyses were conducted for the nine sewersheds. Significant positive linear relationships were found between the wastewater SARS-CoV-2 concentrations and the case numbers reported on the sampling date for all the sites, except Athens (Pearson’s correlation coefficients ranged from 0.38 to 0.89, Figure S2h, Athens’ data excluded from all correlation coefficients presented). Since both viral concentration and case data were highly skewed, violating the assumption of normality underlying the computation of the Pearson correlation coefficient (Figure S1), the non-parametric Spearman rank correlation coefficients were also computed. The correlation coefficients of COVID-19 case counts and the three different SARS-CoV-2 gene concentrations were presented in a heatmap (Figure 3c). The concentrations of all three SARS-CoV-2 genes were significantly, positively correlated with the daily confirmed cases for all sites, except Athens (Spearman’s *r* ranged from 0.48 to 0.87, all *p* < 0.05), among which the N2 gene achieved the best performance, indicated by its highest average Spearman rank correlation coefficients (Spearman’s *r* = 0.70). The robustness of the N2 primer set in quantitative PCR is also shown in other studies, reporting that the primer binding region of the N2 gene is less prone to mutation (52). Therefore, the N2 gene concentrations were used for further statistical analysis.

**Figure 3.**
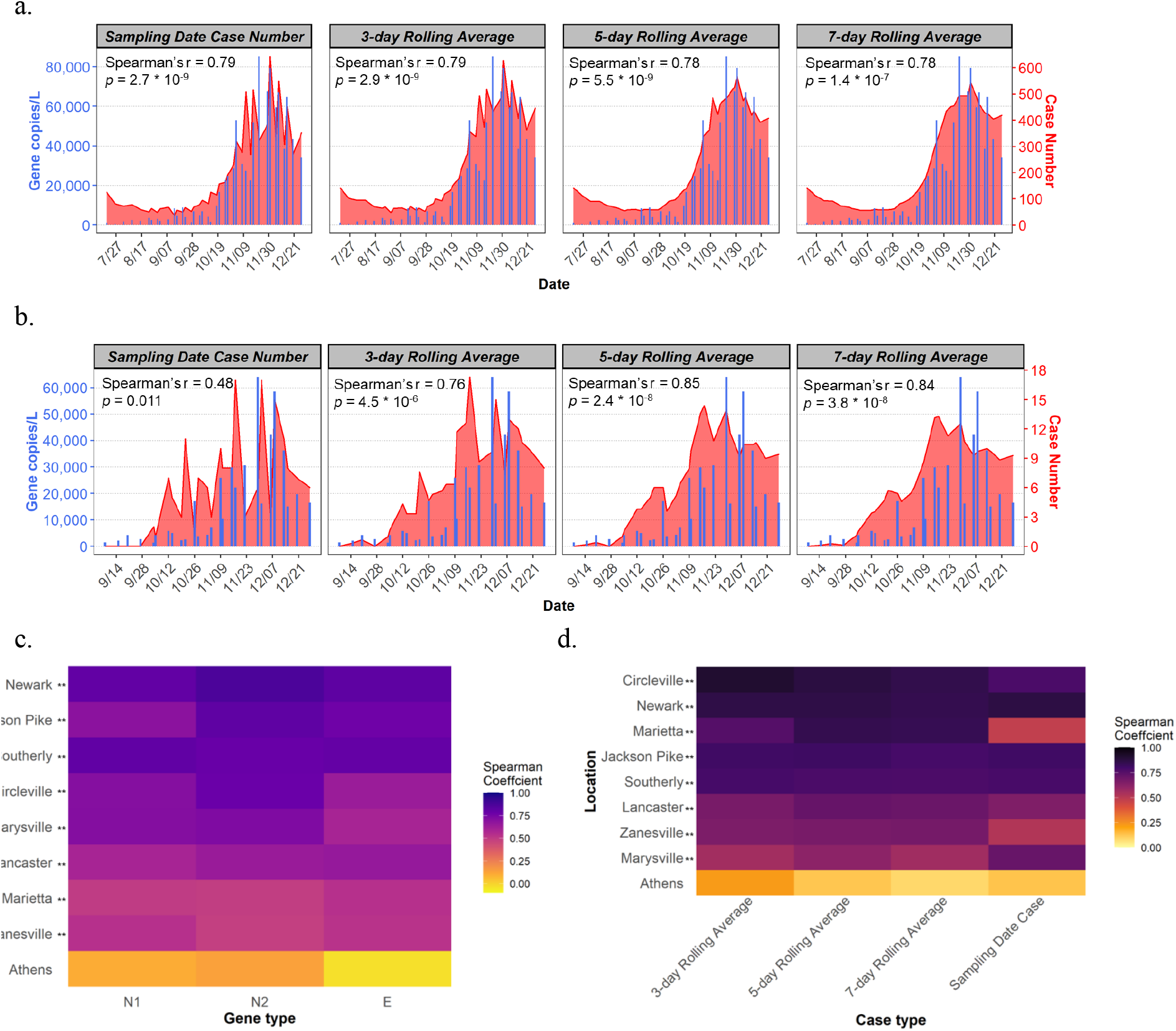
Relationships between SARS-CoV-2 gene concentration in wastewater and new confirmed cases. Overlaid trend plots of SARS-CoV-2 N2 gene concentrations in wastewater and case number of different averaging methods in: a) Southerly sewershed population; b) Marietta sewershed population; c) Spearman correlations of all sites by different genes; and d) Spearman correlations of all sites by case number of different averaging methods. Significant correlations (Spearman) were highlighted with two asterisks and one asterisk for *p-value* < 0.05 and 0.05 < *p-value* < 0.1, respectively.

During an emerging epidemic, a time lag of 3-9 days is typically observed from the onset of symptoms to case reporting, depending on the testing capacity, testing method, care-seeking behavior, and reporting speed (11, 53-54). In addition, the duration of viral shedding in human feces may vary among individuals (4, 55). To overcome these potential uncertainties, rolling averages of the confirmed new case data were calculated in replacement of the daily case numbers in this study. After averaging, the trend of case numbers was smoother and less noisy than the raw case numbers. This was observed in data from both big city, such as Southerly catchment of Columbus (Figure 3a), and small cities like Marietta (Figure 3b). Overlaid trend plots revealed good agreements between wastewater SARS-CoV-2 gene concentrations and new case numbers (Figure S2a-S2g). Pearson (Figure S2i) and Spearman (Figure 3d) correlation coefficients both suggest that the averaged case numbers enhance the extent of relationships between the SARS-CoV-2 gene concentrations and reported COVID-19 cases. The sampling-date case number had the lowest coefficients among the four case types (averaged Spearman’s *r* of all 9 sites = 0.70). The 5-day rolling average of case number showed the highest Spearman rank correlation coefficient (average Spearman’s *r* = 0.77), followed by 7-day (averaged Spearman’s *r* = 0.76) and 3-day averages (average Spearman’s *r* = 0.75). As other studies pointed out that SARS-CoV-2 titers in wastewater may foreshadow the clinical results by 0-4 days, we staggered our WBE trend by 3 days, but found no improvement in the correlations (11, 56-57). It is important to note that the Ohio positive cases are assigned a date of the estimate of disease onset, instead of the test date or the test result date. Considering both sensitivity and precision, the 5-day rolling average of the confirmed new cases was used for understanding the effectiveness of estimating COVID-19 prevalence from wastewater viral concentrations.

The only non-correlating community, Athens, is a small college town with students accounting for 80% population during a university semester. The confirmed cases in Athens showed a different trend compared to other communities, with peak infection rates in mid-September and early October when students returned to campus, declined after that and much lower new case rate in November and December as students left campus by Thanksgiving holiday. Since most college students stay asymptomatic, the discrepancy between the wastewater data and new case data might be explained by an underestimation of the cases in the community, although more testing and analysis would have to be performed to confirm this hypothesis.

Among the nine sewersheds investigated, the wastewater data from four cities correlated remarkably well with the 5-day averaged new case numbers: Circleville WWTP (Spearman’s *r* = 0.88), Newark WWTP (Spearman’s *r* = 0.86), Marietta WWTP (Spearman’s *r* = 0.85), and Jackson Pike WWTP (Spearman’s *r* = 0.81). These findings demonstrate that the wastewater can be used to monitor the dynamic trend of COVID-19 disease in a community, regardless of the population size and magnitude of confirmed cases.

### 3.4. Estimation of COVID-19 cases via wastewater surveillance

Although WBE can provide unbiased samples of the community by aggregating population health information, wastewater is known to have relatively high day-to-day variation in sewage flow and fecal strength (58). To account for the varying estimated fecal load over time and possible dilution due to rain, two human-specific fecal viruses, PMMoV and crAssphage, were quantified from Columbus wastewater to normalize the SARS-CoV-2 concentrations. These two viruses have been used as an internal reference for method validation due to their high persistence in water compared to other bacteria fecal indicators (25). Wastewater from the two Columbus sewersheds are a combined sewer (Table 1), which is more prone to dilution by stormwater events. For each sample analyzed, SARS-CoV-2 RNA concentrations were normalized with the concentrations of PMMoV (mean RNA concentration across the samples: ∼1 × 10^6^ gene copies/L of wastewater) and crAssphage (mean DNA concentration across the samples: ∼1 × 10^8^ gene copies/L of wastewater). Both normalization approaches improved the agreement of the viral concentration and the case counts visually, especially before the change of method (first month) (Figure 4a). According to the Spearman rank correlation, the correlation between the viral titers and 5-day averaged case numbers was only marginally improved by the PMMoV-based normalization but not significant, whereas crAssphage-based normalization led to much weaker correlation.

**Figure 4.**
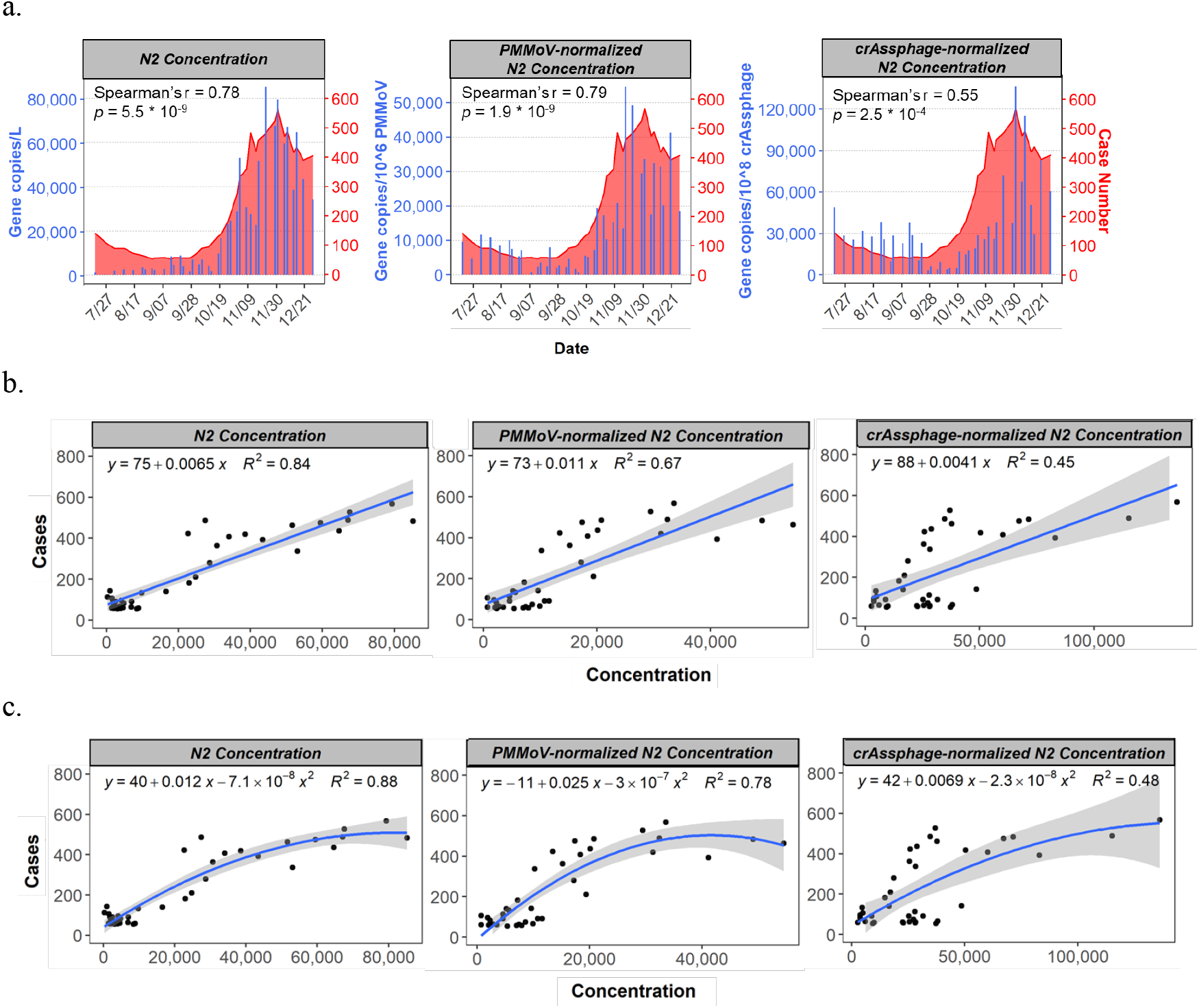
The effect of normalization with human fecal virus indicators on the relationships between SARS-CoV-2 N2 gene concentration in wastewater and 5-day-rolling averages of new confirmed cases. a) Overlaid trend plots of Southerly sewershed population; b) Linear regression model; and c) Quadratic polynomial model.

Our data showed that normalization did not significantly improve wastewater signal correlation with cases during the study period. Although our sites represent a mixture of facilities with separate and combined sewers, during the sampling period we only observed one to three instances per facility where the flows were significantly increased (doubled) compared to the lowest flow recorded during sampling. This primarily occurred in December due to snowmelt (peak of new cases per day), or in August during rain events (new cases close to zero). We ponder that these low frequency precipitation events did not affect the sewer system much, thus normalization of SARS-CoV-2 gene concentrations with human fecal strength did not improve the correlations. In this regard, assumptions can be made that longitudinal wastewater data is robust to the variations in sewage flow and fecal strength when extreme precipitation events are infrequent. Our results agreed with the findings from other studies, which concluded that the PMMoV-normalized SARS-CoV-2 signal had lower background noise and showed the strongest correlation with active cases (57, 59). For future studies where normalization is required, combined markers of viral indicators, solid contents, and other volumetric parameters are recommended (60).

While clinical test results tend to be highly delayed and underestimated, wastewater enables the temporal mapping of the outbreak in a more timely and accurate manner (53). As more and more efforts are put into wastewater surveillance, population health data from wastewater is accumulating worldwide. To make better use of these wastewater information, we tested different models to help estimate COVID-19 prevalence from wastewater viral loads. First, a linear model was built (*R*^*2*^ based on raw N2 gene concentration in Southerly wastewater and 5-day averaged case number = 0.84). Surprisingly, neither normalization approaches enhanced the model (Figure 4b). As mentioned prior, the linear model is not ideal for our dataset and data transformation hardly improved the normality. For a better estimation, polynomial models were built and evaluated. Both quadratic and cubic polynomial models achieved higher *R*^*2*^ than the corresponding linear model. Using the Southerly data as an example, *R*^*2*^ of 0.88 and 0.89, respectively were achieved (Figure 4c). The polynomial models also showed better adherence to normality. However, regression analysis indicated that the cubic polynomial model is not significantly superior to the quadratic model (*p* < 0.05). Based on these results, it can be concluded that the quadratic model gave the best description of the relationship of new COVID-19 cases and viral titers in wastewater, while minimizing overfitting and the violation of normality. Similar to the case in the linear model, normalization did not improve the quadratic model (Figure 4c).

These results demonstrate that dynamic trend of COVID-19 within a community can be well estimated from longitudinal SARS-CoV-2 gene concentrations in wastewater. Polynomial models were built and optimized for the wastewater data from eight of the nine sewersheds. It has been reported that the viral titer in wastewater is more associated with the demographic variables, the household income and medical spend for example, than with population size (57). This may help explain the failure of estimating the disease prevalence with WBE data from Athens. For low-income areas with limited testing capability, it is recommended to take more demographic variables into account when using the WBE as a surveillance tool for the pandemic. Furthermore, recent studies and our preliminary data generated at the early stage of the pandemic (March to May; data now shown) suggested that WBE can be applied to communities of low prevalence (12). As new confirmed cases have been decreasing worldwide due to the implementation of vaccinations, more sensitive and robust methods will perform better in determining SARS-CoV-2 genetic signals in wastewater in the areas with lower COVID-19 prevalence than before.

### 3.5. SARS-CoV-2 variant identification in wastewater

Sequencing of the entire SARS-CoV-2 genome was performed on a subset of wastewater samples. Sequencing results differed among the 8 sites where wastewater samples were obtained over a 3-day period in early January 2021 (Table 2). It was reported that D614G variant dominated the global pandemic over the course of 1 month in March 2020, showing increased infectivity and viral shedding (61). Although all wastewater samples showed nearly 100% D614G mutation, there were variations in the levels of variants associated with the common clades including 20C/G (bearing Q57H), 20B (R203K) and 20A (S194L), with a predominance of clade C at most sites. In addition, there were low levels of other mutations associated with emerging strains, including L452R (associated with the B.1.427/429 strains from California) at Zanesville (10% variant allele frequency (VAF)) and Marietta (7% VAF), as well as N501Y at Jackson Pike (8% VAF). It is notable that wastewater samples from different sewersheds (Jackson Pike and Southerly) in the same city showed unique variant pattern, suggesting the feasibility of deploying wastewater monitoring for rapid detection of emerging variants circulating in each community. The result shows that detected SARS-CoV-2 variants in wastewater agreed well with the sequencing surveillance performed on SARS-CoV-2-positive nasal swabs (Table 2). For example, during January, a N501Y-bearing 20G strain (62) and the B.1.1.429 strain (63) were first detected at low levels in nasopharyngeal swab surveillance samples. Studies have found increased infectivity and transmissibility as well as decreased antibody neutralization in pseudoviruses carrying both L452R and N501Y mutations compared to the D614G alone (64-65). Overall, this study showed the feasibility of identifying circulating SARS-CoV-2 strains in various communities from wastewater.

**Table 2:** Differential detection of SARS-CoV-2 strain-defining mutations by genomic sequencing in wastewater across Ohio over a 3-day period in early January 2021.

| Amino acid change | Nucleotide change | Strain/lineage with mutation | Detection frequency in NP swabs<sup>3</sup> | Variant allele frequency (VAF) |  |  |  |  |  |  |  |
| --- | --- | --- | --- | --- | --- | --- | --- | --- | --- | --- | --- |
|  |  |  |  | Marysville | Jackson Pike | Southerly | Athens | Marietta | Zanesville | Circleville | Lancaster |
| S: D614G<sup>1</sup> | c.1841A>G | B.1, clades A,B,C | 100% | 0.97 | 0.97 | 0.98 | 0.95 | 0.95 | 0.96 | 0.98 | 0.97 |
| ORF3A: Q57H<sup>2</sup> | c.171G>T | B.1, 20C/G clade | 95% | 0.88 | 0.73 | 0.80 | 0.43 | 0.91 | 0.89 | 0.77 | 0.80 |
| N: R203K | c.608G>A | B.1, 20B clade | 3% | nd | nd | nd | 0.19 | nd | nd | nd | nd |
| N: S194L | c.581C>T | B.1, 20A clade | 2% | nd | nd | nd | 0.24 | nd | nd | 0.14 | 0.07 |
| S: N501Y | c.1501A>T | B.1.2/501Y | 4% | nd | 0.08 | nd | nd | nd | nd | nd | nd |
| S: S477N | c.1430G>A | B.1.1.298 & B.1.404 | <1% | nd | nd | 0.16 | nd | nd | nd | 0.08 | nd |
| S: L452R | c.1355T>G | B.1.1.427 & B.1.1.429 | 1% | nd | nd | 0.06 | nd | 0.07 | 0.10 | nd | nd |
| S: P681H | c.2042C>A | B.1.1.222 & B.1.1.7 | 3% | nd | nd | 0.09 | nd | nd | nd | 0.18 | nd |
| N: Q9H | c.27G>T | B.1, 20C subset | 4% | nd | 0.14 | nd | 0.14 | 0.08 | 0.07 | nd | nd |

<sup>1</sup>Allele fraction was well-correlated with other B.1 markers including the 5'UTR C241T, nsp3 p.F106F (c.318C>T) and nsp12 p.P323L (c.968C>T).
<sup>2</sup>Allele fraction was well-correlated with clade 20C/G marker nsp2 p.T85I (c.254C>T).
<sup>3</sup>SARS-CoV-2-positive nasal or nasopharyngeal (NP) swabs collected in January 2021.

### 3.6. Significance of this work

This study demonstrates the effectiveness of wastewater surveillance in COVID-19 trend tracking in various communities. SARS-CoV-2 gene concentrations in wastewater strongly correlated with the COVID-19 cases. This is the first study proposing the use of a quadratic polynomial model to track and predict COVID-19 cases from wastewater surveillance data, which can benefit the communities with limited human testing capability. In the later stage of the pandemic, WBE can help evaluate the effectiveness of vaccination and prioritize the distribution of human testing resources. Moreover, as sequencing results from wastewater samples in early 2021 at a time of new strain emergence shows an agreement with the sequencing results from clinical nasal swab samples, we suggest that the wastewater matrix is an ideal sample for fast tracking variant emergence and transmission within a community.

## Data Availability

Data are available at Ohio Department of Health COVID-19 Dashboard.

https://coronavirus.ohio.gov/wps/portal/gov/covid-19/dashboards/current-trends

## 4. Acknowledgments

This study was supported by funding from Ohio Environmental Protection Agency’s Coronavirus Aid, Relief, and Economic Security (CARES) Act (JL, ZB, SL) and an Interdisciplinary Research Seed Grant from The Ohio State University Infectious Diseases Institute (DJ, HT, JL). Genomic surveillance of nasal swabs was performed under an IRB-approved surveillance protocol (DJ). This research could not be completed without the collaboration with participating wastewater treatment plants in Ohio. We thank Pam Snyder, Sarah Corcoran, and Charlie Andorka at The Ohio State University for their assistance in sample processing and collection and Preeti Pancholi, Sara Koenig and Peter Mohler for logistical support. We appreciate the great support from Rebecca Fugitt at Ohio Department of Health during this study.

## 5. Declaration of Interests

The authors declare no competing interests.

## Notes

### Competing Interest Statement

The authors have declared no competing interest.

### Funding Statement

This study was supported by funding from Ohio Environmental Protection Agency (Coronavirus Aid, Relief, and Economic Security Act) and an Interdisciplinary Research Seed Grant from The Ohio State University Infectious Diseases Institute.

### Author Declarations

Buck IRB and the protocol is 'Clinical Laboratory Investigations and Assay Development for Diagnosis, Monitoring and Outcome Prediction of SARS-CoV-2 Infections and COVID-19 Disease'. Waiver of ethical approval was granted.

## References

1. Hu, B., Guo, H., Zhou, P., Shi, Z.-L., 2021. Characteristics of SARS-CoV-2 and COVID-19. Nat. Rev. Microbiol. 19, 141–154. https://doi.org/10.1038/s41579-020-00459-7

2. World Health Organization (WHO), 2021. WHO Coronavirus Disease (COVID-19) Dashboard [Online]. URL https://covid19.who.int (accessed 3.3.21).

3. Liu, J., Liao, X., Qian, S., Yuan, J., Wang, F., Liu, Y., Wang, Z., Wang, F.-S., Liu, L., Zhang, Z., 2020. Community Transmission of Severe Acute Respiratory Syndrome Coronavirus 2, Shenzhen, China, 2020. Emerg. Infect. Dis. 26, 1320–1323. https://doi.org/10.3201/eid2606.200239

4. He, X., Lau, E.H.Y., Wu, P., Deng, X., Wang, J., Hao, X., Lau, Y.C., Wong, J.Y., Guan, Y., Tan, X., Mo, X., Chen, Y., Liao, B., Chen, W., Hu, F., Zhang, Q., Zhong, M., Wu, Y., Zhao, L., Zhang, F., Cowling, B.J., Li, F., Leung, G.M., 2020. Temporal dynamics in viral shedding and transmissibility of COVID-19. Nat. Med. 26, 672–675. https://doi.org/10.1038/s41591-020-0869-5

5. Hart, O.E., Halden, R.U., 2020. Computational analysis of SARS-CoV-2/COVID-19 surveillance by wastewater-based epidemiology locally and globally: Feasibility, economy, opportunities and challenges. Sci. Total Environ. 730, 138875. https://doi.org/10.1016/j.scitotenv.2020.138875

6. ODH, 2021. COVID-19 Dashborad: Ohio Coronavirus Wastewater Monitoring Network [Online]. URL https://public.tableau.com/views/COVIDWastewater/Dashboard2?:embed=y&:showVizHome=no&:host_url=https%3A%2F%2Fpublic.tableau.com%2F&:embed_code_version=3&:tabs=no&:toolbar=yes&:display_count=yes&:language=en&:loadOrderID=0 (accessed 3.3.21).

7. Sims, N., Kasprzyk-Hordern, B., 2020. Future perspectives of wastewater-based epidemiology: Monitoring infectious disease spread and resistance to the community level. Environ. Int. 139, 105689. https://doi.org/10.1016/j.envint.2020.105689

8. Cheung, K.S., Hung, I.F.N., Chan, P.P.Y., Lung, K.C., Tso, E., Liu, R., Ng, Y.Y., Chu, M.Y., Chung, T.W.H., Tam, A.R., Yip, C.C.Y., Leung, K.-H., Fung, A.Y.-F., Zhang, R.R., Lin, Y., Cheng, H.M., Zhang, A.J.X., To, K.K.W., Chan, K.-H., Yuen, K.-Y., Leung, W.K., 2020. Gastrointestinal Manifestations of SARS-CoV-2 Infection and Virus Load in Fecal Samples From a Hong Kong Cohort: Systematic Review and Meta-analysis. Gastroenterology 159, 81–95. https://doi.org/10.1053/j.gastro.2020.03.065

9. Wang, X., Zheng, J., Guo, L., Yao, H., Wang, L., Xia, X., Zhang, W., 2020. Fecal viral shedding in COVID-19 patients: Clinical significance, viral load dynamics and survival analysis. Virus Res. 289, 198147. https://doi.org/10.1016/j.virusres.2020.198147

10. Medema, G., Heijnen, L., Elsinga, G., Italiaander, R., Brouwer, A., 2020. Presence of SARS-Coronavirus-2 in sewage (preprint). Occupational and Environmental Health. https://doi.org/10.1101/2020.03.29.20045880

11. Nemudryi, A., Nemudraia, A., Wiegand, T., Surya, K., Buyukyoruk, M., Cicha, C., Vanderwood, K.K., Wilkinson, R., Wiedenheft, B., 2020. Temporal Detection and Phylogenetic Assessment of SARS-CoV-2 in Municipal Wastewater. Cell Rep. Med. 1, 100098. https://doi.org/10.1016/j.xcrm.2020.100098

12. Randazzo, W., Truchado, P., Cuevas-Ferrando, E., Simón, P., Allende, A., Sánchez, G., 2020. SARS-CoV-2 RNA in wastewater anticipated COVID-19 occurrence in a low prevalence area. Water Res. 181, 115942. https://doi.org/10.1016/j.watres.2020.115942

13. Wurtzer, S., Marechal, V., Mouchel, J., Maday, Y., Teyssou, R., Richard, E., Almayrac, J., Moulin, L., 2020. Evaluation of lockdown impact on SARS-CoV-2 dynamics through viral genome quantification in Paris wastewaters (preprint). Epidemiology. https://doi.org/10.1101/2020.04.12.20062679

14. Ahmed, W., Angel, N., Edson, J., Bibby, K., Bivins, A., O’Brien, J.W., Choi, P.M., Kitajima, M., Simpson, S.L., Li, J., Tscharke, B., Verhagen, R., Smith, W.J.M., Zaugg, J., Dierens, L., Hugenholtz, P., Thomas, K.V., Mueller, J.F., 2020a. First confirmed detection of SARS-CoV-2 in untreated wastewater in Australia: A proof of concept for the wastewater surveillance of COVID-19 in the community. Sci. Total Environ. 728, 138764. https://doi.org/10.1016/j.scitotenv.2020.138764

15. Lodder, W., Husman, A.M. de R., 2020. SARS-CoV-2 in wastewater: potential health risk, but also data source. Lancet Gastroenterol. Hepatol. 5, 533–534. https://doi.org/10.1016/S2468-1253(20)30087-X

16. Rosa, G.L., Iaconelli, M., Mancini, P., Ferraro, G.B., Bonadonna, L., Lucentini, L., Suffredini, E., n.d. FIRST DETECTION OF SARS-COV-2 IN UNTREATED WASTEWATERS IN ITALY 17.

17. Wu, F., Zhang, J., Xiao, A., Gu, X., Lee, W.L., Armas, F., Kauffman, K., Hanage, W., Matus, M., Ghaeli, N., Endo, N., Duvallet, C., Poyet, M., Moniz, K., Washburne, A.D., Erickson, T.B., Chai, P.R., Thompson, J., Alm, E.J., 2020b. SARS-CoV-2 Titers in Wastewater Are Higher than Expected from Clinically Confirmed Cases. mSystems 5. https://doi.org/10.1128/mSystems.00614-20

18. Kevin, L., 2020. Back log of test results causes delayed COVID-19 numbers reported by ODH [Online]. URL https://www.10tv.com/article/news/health/coronavirus/back-log-of-test-results-causes-delayed-covid-19-numbers-reported-by-odh/530-2e377780-687b-4ee2-b7fb-8f9991bb15d0 (accessed 3.11.21).

19. Richterich, P., 2020. Severe underestimation of COVID-19 case numbers: effect of epidemic growth rate and test restrictions (preprint). Infectious Diseases (except HIV/AIDS). https://doi.org/10.1101/2020.04.13.20064220

20. Ahmed, W., Tscharke, B., Bertsch, P.M., Bibby, K., Bivins, A., Choi, P., Clarke, L., Dwyer, J., Edson, J., Nguyen, T.M.H., O’Brien, J.W., Simpson, S.L., Sherman, P., Thomas, K.V., Verhagen, R., Zaugg, J., Mueller, J.F., 2021. SARS-CoV-2 RNA monitoring in wastewater as a potential early warning system for COVID-19 transmission in the community: A temporal case study. Sci. Total Environ. 761, 144216. https://doi.org/10.1016/j.scitotenv.2020.144216

21. Fontenele, R.S., Kraberger, S., Hadfield, J., Driver, E.M., Bowes, D., Holland, L.A., Faleye, T.O.C., Adhikari, S., Kumar, R., Inchausti, R., Holmes, W.K., Deitrick, S., Brown, P., Duty, D., Smith, T., Bhatnagar, A., Yeager, R.A., Holm, R.H. Reitzenstein, N.H. von, Wheeler, E., Dixon, K., Constantine, T., Wilson, M.A., Lim, E.S., Jiang, X., Halden, R.U., Scotch, M., Varsani, A., 2021. High-throughput sequencing of SARS-CoV-2 in wastewater provides insights into circulating variants. medRxiv 2021.01.22.21250320. https://doi.org/10.1101/2021.01.22.21250320

22. Jahn, K., Dreifuss, D., Topolsky, I., Kull, A., Ganesanandamoorthy, P., Fernandez-Cassi, X., Bänziger, C., Stachler, E., Fuhrmann, L., Jablonski, K.P., Chen, C., Aquino, C., Stadler, T., Ort, C., Kohn, T., Julian, T.R., Beerenwinkel, N., 2021. Detection of SARS-CoV-2 variants in Switzerland by genomic analysis of wastewater samples. medRxiv 2021.01.08.21249379. https://doi.org/10.1101/2021.01.08.21249379

23. Martin, J., Klapsa, D., Wilton, T., Zambon, M., Bentley, E., Bujaki, E., Fritzsche, M., Mate, R., Majumdar, M., 2020. Tracking SARS-CoV-2 in Sewage: Evidence of Changes in Virus Variant Predominance during COVID-19 Pandemic. Viruses 12, 1144. https://doi.org/10.3390/v12101144

24. Daughton, C.G., 2012. Using biomarkers in sewage to monitor community-wide human health: Isoprostanes as conceptual prototype. Sci. Total Environ. 424, 16–38. https://doi.org/10.1016/j.scitotenv.2012.02.038

25. Greaves, J., Stone, D., Wu, Z., Bibby, K., 2020. Persistence of emerging viral fecal indicators in large-scale freshwater mesocosms. Water Res. X 9, 100067. https://doi.org/10.1016/j.wroa.2020.100067

26. Hamza, I.A., Jurzik, L., Überla, K., Wilhelm, M., 2011. Evaluation of pepper mild mottle virus, human picobirnavirus and Torque teno virus as indicators of fecal contamination in river water. Water Res. 45, 1358–1368. https://doi.org/10.1016/j.watres.2010.10.021

27. Malla, B., Ghaju Shrestha, R., Tandukar, S., Sherchand, J.B., Haramoto, E., 2019. Performance Evaluation of Human-Specific Viral Markers and Application of Pepper Mild Mottle Virus and CrAssphage to Environmental Water Samples as Fecal Pollution Markers in the Kathmandu Valley, Nepal. Food Environ. Virol. 11, 274–287. https://doi.org/10.1007/s12560-019-09389-x

28. Symonds, E.M., Nguyen, K.H., Harwood, V.J., Breitbart, M., 2018. Pepper mild mottle virus: A plant pathogen with a greater purpose in (waste)water treatment development and public health management. Water Res. 144, 1–12. https://doi.org/10.1016/j.watres.2018.06.066

29. U.S. Department of Commerce, 2019. U.S. Census Bureau QuickFacts: United States [Online]. URL https://www.census.gov/quickfacts/fact/table/US/PST045219 (accessed 3.9.21).

30. Bennett, H.B., O’Dell, H.D., Norton, G., Shin, G., Hsu, F.-C., Meschke, J.S., 2010. Evaluation of a novel electropositive filter for the concentration of viruses from diverse water matrices. Water Sci. Technol. J. Int. Assoc. Water Pollut. Res. 61, 317–322. https://doi.org/10.2166/wst.2010.819

31. Innovaprep, 2020. Concentrating Pipette Select: Wastewater Application Note Revision B (p. 2) [Online]. URL https://uploads-ssl.webflow.com/57aa3257c3e841c509f276e2/5f888d1b3bddf35ae661965c_CONCENTRATINGPIPETTESELECT%20WASTEWATER%20APPLICATION%20NOTE%201.17.03%20PM-compressed.pdf (accessed 3.3.21).

32. Bae, J., Schwab, K.J., 2008. Evaluation of Murine Norovirus, Feline Calicivirus, Poliovirus, and MS2 as Surrogates for Human Norovirus in a Model of Viral Persistence in Surface Water and Groundwater. Appl. Environ. Microbiol. 74, 477–484. https://doi.org/10.1128/AEM.02095-06

33. Cimolai, N., 2020. Environmental and decontamination issues for human coronaviruses and their potential surrogates. J. Med. Virol. 92, 2498–2510. https://doi.org/10.1002/jmv.26170

34. Hirotsu, Y., Mochizuki, H., Omata, M., 2020. Double-Quencher Probes Improved the Detection Sensitivity of Severe Acute Respiratory Syndrome Coronavirus 2 (SARS-CoV-2) by One-Step RT-PCR. medRxiv 2020.03.17.20037903. https://doi.org/10.1101/2020.03.17.20037903

35. Jung, Y.J., Park, G.-S., Moon, J.H., Ku, K., Beak, S.-H., Kim, S., Park, E.C., Park, D., Lee, J.-H., Byeon, C.W., Lee, J.J., Maeng, J.-S., Kim, S.J., Kim, S.I., Kim, B.-T., Lee, M.J., Kim, H.G., 2020. Comparative analysis of primer-probe sets for the laboratory confirmation of SARS-CoV-2. bioRxiv 2020.02.25.964775. https://doi.org/10.1101/2020.02.25.964775

36. Corman, V.M., Landt, O., Kaiser, M., Molenkamp, R., Meijer, A., Chu, D.K., Bleicker, T., Brünink, S., Schneider, J., Schmidt, M.L., Mulders, D.G., Haagmans, B.L., Veer, B. van der, Brink, S. van den, Wijsman, L., Goderski, G., Romette, J.-L., Ellis, J., Zambon, M., Peiris, M., Goossens, H., Reusken, C., Koopmans, M.P., Drosten, C., 2020. Detection of 2019 novel coronavirus (2019-nCoV) by real-time RT-PCR. Eurosurveillance 25, 2000045. https://doi.org/10.2807/1560-7917.ES.2020.25.3.2000045

37. Dare, R.K., Fry, A.M., Chittaganpitch, M., Sawanpanyalert, P., Olsen, S.J., Erdman, D.D., 2007. Human Coronavirus Infections in Rural Thailand: A Comprehensive Study Using Real-Time Reverse-Transcription Polymerase Chain Reaction Assays. J. Infect. Dis. 196, 1321–1328. https://doi.org/10.1086/521308

38. Decaro, N., Elia, G., Campolo, M., Desario, C., Mari, V., Radogna, A., Colaianni, M.L., Cirone, F., Tempesta, M., Buonavoglia, C., 2008. Detection of bovine coronavirus using a TaqMan-based real-time RT-PCR assay. J. Virol. Methods 151, 167–171. https://doi.org/10.1016/j.jviromet.2008.05.016

39. Haramoto, E., Kitajima, M., Kishida, N., Konno, Y., Katayama, H., Asami, M., Akiba, M., 2013. Occurrence of Pepper Mild Mottle Virus in Drinking Water Sources in Japan. Appl. Environ. Microbiol. 79, 7413–7418. https://doi.org/10.1128/AEM.02354-13

40. Ogorzaly, L., Gantzer, C., 2006. Development of real-time RT-PCR methods for specific detection of F-specific RNA bacteriophage genogroups: Application to urban raw wastewater. J. Virol. Methods 138, 131–139. https://doi.org/10.1016/j.jviromet.2006.08.004

41. Stachler, E., Kelty, C., Sivaganesan, M., Li, X., Bibby, K., Shanks, O.C., 2017. Quantitative CrAssphage PCR Assays for Human Fecal Pollution Measurement. Environ. Sci. Technol. 51, 9146–9154. https://doi.org/10.1021/acs.est.7b02703

42. Johnson, D.R., Lee, P.K.H., Holmes, V.F., Alvarez-Cohen, L., 2005. An internal reference technique for accurately quantifying specific mRNAs by real-time PCR with application to the tceA reductive dehalogenase gene. Appl. Environ. Microbiol. 71, 3866–3871. https://doi.org/10.1128/AEM.71.7.3866-3871.2005

43. Rambaut, A., Holmes, E.C., O’Toole, Á., Hill, V., McCrone, J.T., Ruis, C., du Plessis, L., Pybus, O.G., 2020. A dynamic nomenclature proposal for SARS-CoV-2 lineages to assist genomic epidemiology. Nat. Microbiol. 5, 1403–1407. https://doi.org/10.1038/s41564-020-0770-5

44. Bedford, T., Hodcroft, E., Neher, R., n.d. Updated Nextstrain SARS-CoV-2 clade naming strategy [Online]. Updat. Nextstrain SARS-CoV-2 Clade Naming Strategy. URL https://nextstrain.org//blog/2021-01-06-updated-SARS-CoV-2-clade-naming (accessed 4.14.21).

45. M. Pecson B., Darby, E., N. Haas, C. M. Amha Y., Bartolo, M., Danielson, R., Dearborn, Y., Giovanni, G.D., Ferguson, C., Fevig, S., Gaddis, E., Gray, D., Lukasik, G., Mull, B., Olivas, L., Olivieri, A., Qu, Y., Consortium, S.-C.-2 I., 2021. Reproducibility and sensitivity of 36 methods to quantify the SARS-CoV-2 genetic signal in raw wastewater: findings from an interlaboratory methods evaluation in the U.S. Environ. Sci. Water Res. Technol. https://doi.org/10.1039/D0EW00946F

46. Dawson, D.J., Paish, A., Staffell, L.M., Seymour, I.J., Appleton, H., 2005. Survival of viruses on fresh produce, using MS2 as a surrogate for norovirus. J. Appl. Microbiol. 98, 203–209. https://doi.org/10.1111/j.1365-2672.2004.02439.x

47. Lin, K., Marr, L.C., 2017. Aerosolization of Ebola Virus Surrogates in Wastewater Systems. Environ. Sci. Technol. 51, 2669–2675. https://doi.org/10.1021/acs.est.6b04846

48. Ahmed, W., Bertsch, P.M., Bivins, A., Bibby, K., Farkas, K., Gathercole, A., Haramoto, E., Gyawali, P., Korajkic, A., McMinn, B.R., Mueller, J.F., Simpson, S.L., Smith, W.J.M., Symonds, E.M., Thomas, K.V., Verhagen, R., Kitajima, M., 2020b. Comparison of virus concentration methods for the RT-qPCR-based recovery of murine hepatitis virus, a surrogate for SARS-CoV-2 from untreated wastewater. Sci. Total Environ. 739, 139960. https://doi.org/10.1016/j.scitotenv.2020.139960

49. Ye, Y., Ellenberg, R.M., Graham, K.E., Wigginton, K.R., 2016. Survivability, Partitioning, and Recovery of Enveloped Viruses in Untreated Municipal Wastewater. Environ. Sci. Technol. 50, 5077–5085. https://doi.org/10.1021/acs.est.6b00876

50. Sedlak, R.H., Kuypers, J., Jerome, K.R., 2014. A multiplexed droplet digital PCR assay performs better than qPCR on inhibition prone samples. Diagn. Microbiol. Infect. Dis. 80, 285–286. https://doi.org/10.1016/j.diagmicrobio.2014.09.004

51. CDC, 2020a. COVID Data Tracker [Online]. Cent. Dis. Control Prev. URL https://covid.cdc.gov/covid-data-tracker (accessed 3.10.21).

52. Rahman, M.S., Islam, M.R., Alam, A.S.M.R.U., Islam, I., Hoque, M.N., Akter, S., Rahaman, M.M., Sultana, M., Hossain, M.A., 2021. Evolutionary dynamics of SARS-CoV-2 nucleocapsid protein and its consequences. J. Med. Virol. 93, 2177–2195. https://doi.org/10.1002/jmv.26626

53. Garg, S., Kim, L., Whitaker, M., O’Halloran, A., Cummings, C., Holstein, R., Prill, M., Chai, S.J., Kirley, P.D., Alden, N.B., Kawasaki, B., Yousey-Hindes, K., Niccolai, L., Anderson, E.J., Openo, K.P., Weigel, A., Monroe, M.L., Ryan, P., Henderson, J., Kim, S., Como-Sabetti, K., Lynfield, R., Sosin, D., Torres, S., Muse, A., Bennett, N.M., Billing, L., Sutton, M., West, N., Schaffner, W., Talbot, H.K., Aquino, C., George, A., Budd, A., Brammer, L., Langley, G., Hall, A.J., Fry, A., 2020. Hospitalization Rates and Characteristics of Patients Hospitalized with Laboratory-Confirmed Coronavirus Disease 2019 — COVID-NET, 14 States, March 1–30, 2020. Morb. Mortal. Wkly. Rep. 69, 458– 464. https://doi.org/10.15585/mmwr.mm6915e3

54. Silverman, J.D., Hupert, N., Washburne, A.D., 2020. Using influenza surveillance networks to estimate state-specific prevalence of SARS-CoV-2 in the United States. Sci. Transl. Med. 12. https://doi.org/10.1126/scitranslmed.abc1126

55. Xiao, F., Tang, M., Zheng, X., Liu, Y., Li, X., Shan, H., 2020. Evidence for Gastrointestinal Infection of SARS-CoV-2. Gastroenterology 158, 1831-1833.e3. https://doi.org/10.1053/j.gastro.2020.02.055

56. Peccia, J., Zulli, A., Brackney, D.E., Grubaugh, N.D., Kaplan, E.H., Casanovas-Massana, A., Ko, A.I., Malik, A.A., Wang, D., Wang, M., Warren, J.L., Weinberger, D.M., Arnold, W., Omer, S.B., 2020a. Measurement of SARS-CoV-2 RNA in wastewater tracks community infection dynamics. Nat. Biotechnol. 38, 1164–1167. https://doi.org/10.1038/s41587-020-0684-z

57. Wu, F., Xiao, A., Zhang, J., Moniz, K., Endo, N., Armas, F., Bonneau, R., Brown, M.A., Bushman, M., Chai, P.R., Duvallet, C., Erickson, T.B., Foppe, K., Ghaeli, N., Gu, X., Hanage, W.P., Huang, K.H., Lee, W.L., Matus, M., McElroy, K.A., Nagler, J., Rhode, S.F., Santillana, M., Tucker, J.A., Wuertz, S., Zhao, S., Thompson, J., Alm, E.J., 2020a. SARS-CoV-2 titers in wastewater foreshadow dynamics and clinical presentation of new COVID-19 cases. medRxiv. https://doi.org/10.1101/2020.06.15.20117747

58. Peccia, J., Zulli, A., Brackney, D.E., Grubaugh, N.D., Kaplan, E.H., Casanovas-Massana, A., Ko, A.I., Malik, A.A., Wang, D., Wang, M., Warren, J.L., Weinberger, D.M., Omer, S.B., 2020b. SARS-CoV-2 RNA concentrations in primary municipal sewage sludge as a leading indicator of COVID-19 outbreak dynamics. medRxiv 2020.05.19.20105999. https://doi.org/10.1101/2020.05.19.20105999

59. D’Aoust, P.M., Mercier, E., Montpetit, D., Jia, J.-J., Alexandrov, I., Neault, N., Baig, A.T., Mayne, J., Zhang, X., Alain, T., Langlois, M.-A., Servos, M.R., MacKenzie, M., Figeys, D., MacKenzie, A.E., Graber, T.E., Delatolla, R., 2021. Quantitative analysis of SARS-CoV-2 RNA from wastewater solids in communities with low COVID-19 incidence and prevalence. Water Res. 188, 116560. https://doi.org/10.1016/j.watres.2020.116560

60. Neault, N., Baig, A.T., Graber, T.E., D’Aoust, P.M., Mercier, E., Alexandrov, I., Crosby, D., Baird, S., Mayne, J., Pounds, T., MacKenzie, M., Figeys, D., MacKenzie, A., Delatolla, R., 2020. SARS-CoV-2 Protein in Wastewater Mirrors COVID-19 Prevalence. medRxiv 2020.09.01.20185280. https://doi.org/10.1101/2020.09.01.20185280

61. Korber, B., Fischer, W.M., Gnanakaran, S., Yoon, H., Theiler, J., Abfalterer, W., Hengartner, N., Giorgi, E.E., Bhattacharya, T., Foley, B., Hastie, K.M., Parker, M.D., Partridge, D.G., Evans, C.M., Freeman, T.M., de Silva, T.I., Angyal, A., Brown, R.L., Carrilero, L., Green, L.R., Groves, D.C., Johnson, K.J., Keeley, A.J., Lindsey, B.B., Parsons, P.J., Raza, M., Rowland-Jones, S., Smith, N., Tucker, R.M., Wang, D., Wyles, M.D., McDanal, C., Perez, L.G., Tang, H., Moon-Walker, A., Whelan, S.P., LaBranche, C.C., Saphire, E.O., Montefiori, D.C., 2020. Tracking Changes in SARS-CoV-2 Spike: Evidence that D614G Increases Infectivity of the COVID-19 Virus. Cell 182, 812-827.e19. https://doi.org/10.1016/j.cell.2020.06.043

62. Tu, H., Avenarius, M.R., Kubatko, L., Hunt, M., Pan, X., Ru, P., Garee, J., Thomas, K., Mohler, P., Pancholi, P., Jones, D., 2021. Distinct Patterns of Emergence of SARS-CoV-2 Spike Variants including N501Y in Clinical Samples in Columbus Ohio. bioRxiv 2021.01.12.426407. https://doi.org/10.1101/2021.01.12.426407

63. CDC, 2020b. Cases, Data, and Surveillance [Online]. Cent. Dis. Control Prev. URL https://www.cdc.gov/coronavirus/2019-ncov/cases-updates/variant-surveillance/variant-info.html (accessed 3.29.21).

64. Deng, X., Garcia-Knight, M.A., Khalid, M.M., Servellita, V., Wang, C., Morris, M.K., Sotomayor-González, A., Glasner, D.R., Reyes, K.R., Gliwa, A.S., Reddy, N.P., Martin, C.S.S., Federman, S., Cheng, J., Balcerek, J., Taylor, J., Streithorst, J.A., Miller, S., Kumar, G.R., Sreekumar, B., Chen, P.-Y., Schulze-Gahmen, U., Taha, T.Y., Hayashi, J., Simoneau, C.R., McMahon, S., Lidsky, P.V., Xiao, Y., Hemarajata, P., Green, N.M., Espinosa, A., Kath, C., Haw, M., Bell, J., Hacker, J.K., Hanson, C., Wadford, D.A., Anaya, C., Ferguson, D., Lareau, L.F., Frankino, P.A., Shivram, H., Wyman, S.K., Ott, M., Andino, R., Chiu, C.Y., 2021. Transmission, infectivity, and antibody neutralization of an emerging SARS-CoV-2 variant in California carrying a L452R spike protein mutation. medRxiv 2021.03.07.21252647. https://doi.org/10.1101/2021.03.07.21252647

65. Xie, X., Liu, Y., Liu, J., Zhang, X., Zou, J., Fontes-Garfias, C.R., Xia, H., Swanson, K.A., Cutler, M., Cooper, D., Menachery, V.D., Weaver, S.C., Dormitzer, P.R., Shi, P.-Y., 2021. Neutralization of SARS-CoV-2 spike 69/70 deletion, E484K and N501Y variants by BNT162b2 vaccine-elicited sera. Nat. Med. 1–2. https://doi.org/10.1038/s41591-021-01270-4

